# Three multimodal large language models fail at clinically actionable breast pathology in three different directions

**DOI:** 10.64898/2026.06.18.26355928

**Authors:** Sun-Young Jun, Seonhui Kim, Young-Joon Kang

**Affiliations:** Department of Pathology, Incheon St. Mary’s Hospital, College of Medicine, The Catholic University of Korea, Seoul, Republic of Korea; Department of Surgery, Incheon St. Mary’s Hospital, College of Medicine, The Catholic University of Korea, Seoul, Republic of Korea

**Keywords:** Breast Neoplasms, Large Language Models, Artificial Intelligence, Image Interpretation, Computer-Assisted, Telepathology, Observer Variation, Neoplasm Grading, Developing Countries

## Abstract

**Background:** Breast cancer treatment depends on histopathological features, such as grade and receptor-defined subtype; however, specialist pathologist access is constrained when the workforce is limited. Commercial multimodal large language models (MLLMs) accept hematoxylin and eosin (H&E) image tiles through paid interfaces without local hardware or fine-tuning. However, prior pathology evaluations addressed only coarse tasks. Whether they reach treatment-determining accuracy and whether vendors agree remain unclear.

**Methods:** We aimed to evaluate three vendor-designated flagship MLLMs (Claude Sonnet 4.6, Gemini 2.5 Pro, GPT-5.5) in 427 invasive breast cancer cases. Each case went to all three with identical H&E tiles and prompts, and the subtype was inferred in the second call. The reference was an institutional sign-out report of an immunohistochemistry-derived subtype. We calculated the concordance, sensitivity, specificity, Cohen’s kappa, and pairwise McNemar and Bowker tests.

**Findings:** Claude ranked highest by raw histologic-type concordance but lowest by kappa, classifying all 23 lobular and seven micropapillary carcinomas as invasive breast carcinoma of no special type. The models anchored the Nottingham grade to three modal grades. None of the models reliably identified human epidermal growth factor receptor 2-positive disease. The failure direction was vendor-specific: Claude and GPT-5.5 were under-detected, whereas Gemini was over-called. Twelve prompt variants (4,056 calls) did not recover sensitivity.

**Interpretation:** No current commercial MLLM reaches deployment-ready accuracy for any treatment-determining feature of breast pathology. As each vendor fails in its own fixed direction, changing vendors alters the type of error rather than removing it; therefore, the value of these models is assistive rather than autonomous. At USD 0.20–0.50 per case, they may serve as supervised draft generators that leave the diagnosis with the pathologist.

**Research in context:** *Evidence before this study:* We searched PubMed and Embase from database inception to January 10, 2026, without language restriction, using combinations of the terms “large language model,” “multimodal,” “GPT,” “Gemini,” “Claude,” “foundation model,” “breast cancer,” “pathology,” “histopathology,” “whole-slide image,” and “diagnostic accuracy.” We also screened the reference lists of retrieved articles. Task-specific deep-learning models and pathology foundation models pretrained on large slide collections achieve strong performance on individual breast pathology tasks such as Nottingham grading and receptor-status prediction, but require local GPU infrastructure, curated training data, and deployment expertise. Evaluations of general-purpose commercially available multimodal large language models (MLLMs) in pathology were limited to coarse tasks, such as tissue-type classification and metastasis, and were typically confined to a single model. We found no studies directly comparing current flagship commercial MLLMs on clinically relevant, treatment-determining breast pathology tasks, and none reporting whether vendors agree or whether the choice of vendor changes the diagnosis. The available evidence was limited by single-model designs, task-narrow datasets, and reliance on raw accuracy without chance-corrected agreement.

*Added value of this study:* In this paired, single-center retrospective study of 427 invasive breast cancers, we evaluated three vendor-designated flagship commercial MLLMs (Claude Sonnet 4.6, Gemini 2.5 Pro, GPT-5.5) on identical H&E tiles and prompts against an expert pathologist reference, across treatment-determining features. No model reached deployment-ready accuracy for any feature. Raw concordance and chance-corrected agreement ranked the models in opposite order, so a decision based on raw accuracy alone would have selected the least discriminating model. Each model demonstrated a consistent error pattern, tending either toward under-detection or over-call. Consequently, changing vendors altered the type of diagnostic error rather than removing it. Twelve prompt variants across 4,056 calls, a reasoning-effort escalation, and an intra-vendor version upgrade did not change the failure direction, indicating a vendor-specific prior rather than a prompt-engineering artifact. To our knowledge, this is the first head-to-head, chance-corrected comparison of commercial MLLMs at the level of treatment-determining breast pathology.

*Implications of all the available evidence:* Taken together with previous evidence, our findings indicate that current commercial MLLMs are not ready for autonomous interpretation of breast pathology and should not be used as primary readers without pathologist oversight. Their value, if any, is assistive. Given their relatively low per-case cost, these systems may be useful for generating supervised draft reports where pathologist workload is the primary constraint, provided that digital pathology infrastructure and immunohistochemical testing are available and that final diagnostic responsibility remains with a qualified pathologist. Because the failure direction is vendor-specific, deployment requires vendor-aware caveats and item-level, chance-corrected evaluation rather than raw accuracy. Improving performance will likely require incorporation of domain-specific knowledge through approaches such as in-context reference examples, retrieval against a curated atlas, or handoff to a domain-trained model. Future development should be supported by immunohistochemistry-grounded datasets and prospective validation in the target setting.

## Introduction

Breast cancer is the most commonly diagnosed cancer in women worldwide.^1^ The burden is shifting toward low- and middle-income countries (LMICs), where the 5-year survival is lower than that in high-income settings,^2,3^ largely owing to late-stage presentation and limited diagnostic infrastructure.^3^

The histopathology of resected specimens remains the reference standard for breast cancer diagnosis and a determinant of subsequent treatment selection.^4,5^ However, access to pathology services is uneven. In a Lancet series on pathology and laboratory medicine, sub-Saharan Africa had fewer than one pathologist per 500,000 people in most countries, with parallel deficits in trained technologists, immunohistochemistry (IHC) capacity, and quality assurance.^6,7^ Cancer care in sub-Saharan Africa depends on pathology services that remain underdeveloped relative to need.^8^ Even high-income settings show a declining capacity; the active US pathology workforce decreased by 17.5% between 2007 and 2017, whereas the diagnostic workload per pathologist increased by 41.7%.^9^ The World Health Organization (WHO) Global Breast Cancer Initiative has identified diagnostic capacity as one of the three priority pillars for reducing breast cancer mortality in LMICs.^3^

The current alternatives impose deployment constraints. Telepathology has been validated for cancer triage in rural settings. However, it requires an available remote pathologist, a static or whole-slide imaging workflow, and reliable connectivity.^10^ Computational pathology approaches, including weakly supervised deep learning^11,12^ and pathology-domain foundation models pretrained on large-scale whole-slide-image or image-caption datasets,^13–15^ achieve strong performance on downstream tasks but require a local graphics processing unit (GPU) infrastructure, model deployment expertise, and curated training data, restricting access to well-resourced centers. Vision-language assistants, such as PathChat, add image-grounded conversational reasoning but follow the same domain-training paradigm.^16^

Commercial multimodal large language models (MLLMs) accept image inputs along with structured prompts and return free-text or JavaScript Object Notation (JSON) outputs through an application programming interface (API) without local model training, GPU hardware, or domain fine-tuning. From a deployment perspective, the requirements are an Internet connection and an API account, and the technical barrier is lower than that of the foundation model deployment or telepathology with on-call expertise. Three vendors now offer general availability flagship multimodal models, but evaluations of these models in pathology have been limited to coarse tasks,^17,18^ not the treatment-determining features that guide therapy. Whether any reaches the accuracy required for a treatment-determining reading, whether the models agree with one another, and whether the choice of vendor alters the resulting diagnosis have not yet been examined.

We conducted a paired head-to-head concordance study of three flagship commercial MLLMs (Claude Sonnet 4.6, Gemini 2.5 Pro, GPT-5.5) on hematoxylin and eosin (H&E) images from 427 invasive breast cancer cases, with the institutional pathology report as a reference. We assessed item-level concordance for treatment-determining features, compared raw and chance-corrected agreements, characterized the direction of each model’s errors, and tested whether the prompt redesign alters these patterns.

## Methods

### Study design

This was a single-center, retrospective concordance study comparing the diagnostic output of three commercial MLLMs with an expert pathologist’s reference to invasive breast cancer. Specimens were collected at a Korean academic hospital and were predominantly surgical resections, with a small subset of core-needle biopsy or frozen-section material. Reporting followed the STAndards for the Reporting of Diagnostic accuracy - Artificial Intelligence (STARD-AI), where applicable. The workflow of this study is summarized in Figure 1.

**Figure 1.**
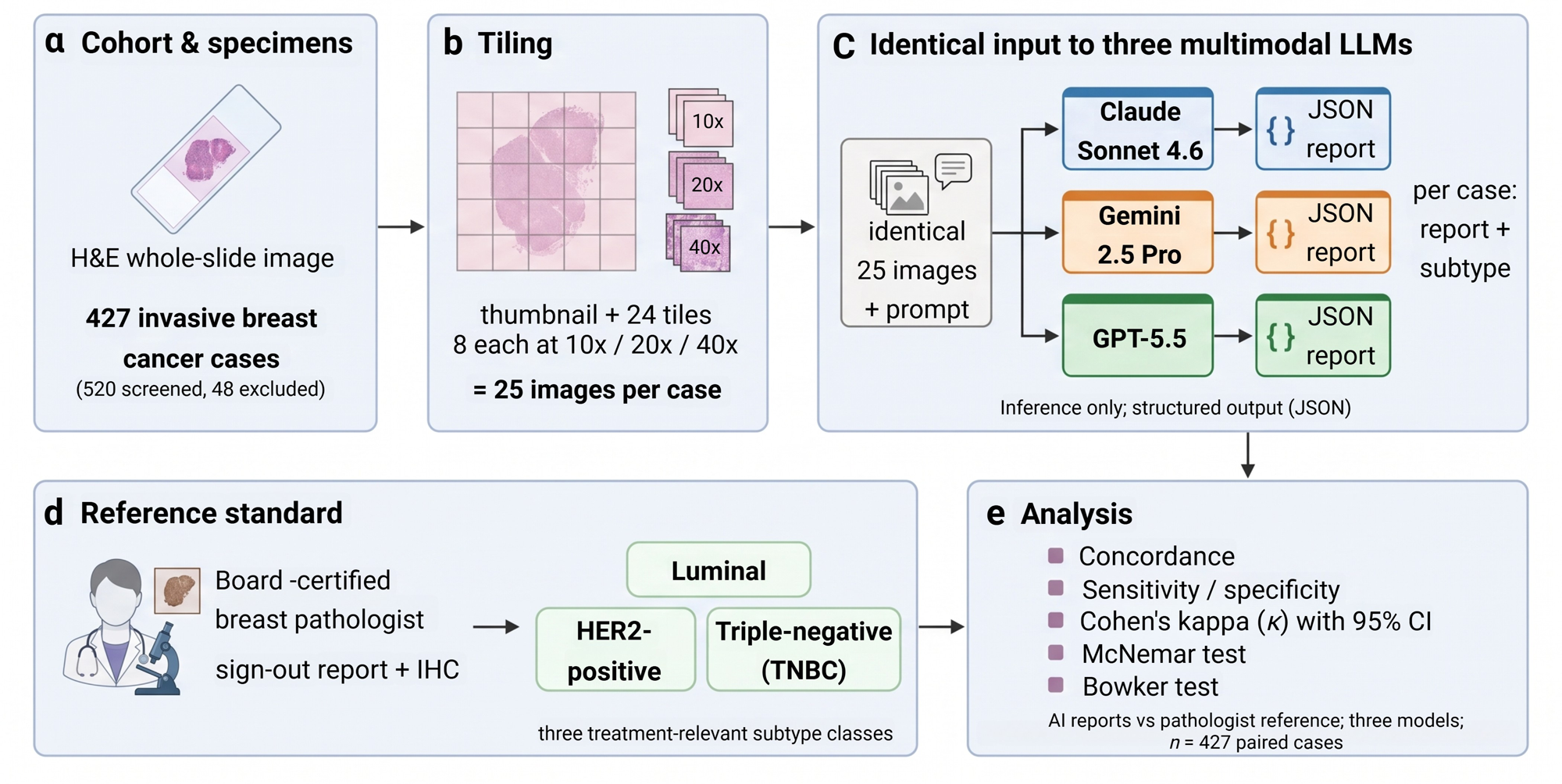
Study workflow (Methods overview)

### Cohort and case selection

Of the 520 available breast whole-slide images (WSIs), 31 lacked a matched clinical pathology report, leaving 489. Cases were excluded if the report described in-situ-only disease, recurrence, distant metastasis, a lymph node-only slide, postoperative re-excision, or external consultation material. When ancillary core-needle biopsy or frozen-section report and a resection report were available, the resection report served as the reference. We excluded 48 reports based on these criteria and removed 10 duplicate bilateral specimens via per-patient deduplication, leaving 431. A further two cases lacked usable WSI, and two failed model inferences, leaving 427 cases analyzed across all three models. The molecular subtype was evaluated in 414 cases using classifiable IHC.

### Hematoxylin and eosin whole-slide image processing

Each case contributed a single representative WSI of the primary invasive tumor in the Aperio SVS format and was processed using OpenSlide. From each slide, an automated routine extracted a low-magnification thumbnail of the entire section (1536×1536 px), together with eight tiles at three magnifications (10×, 20×, and 40× equivalent), giving 25 images per case. Tile coordinates favored regions of high cellularity, intermediate stromal density, and peripheral tumor–stroma interface, and the same coordinates were reused for all three models so that every model received an identical image set. The pipeline operates on a single central processing unit and does not require a GPU.

### Models and configuration

We evaluated the model designated by each vendor as its current general availability flagship multimodal system as of May 2026: Anthropic Claude Sonnet 4.6 (released 2026-02-17), Google Gemini 2.5 Pro (general availability release 2025-06-17), and OpenAI GPT-5.5 (released 2026-04-23). Vendor defaults were kept close to neutral: temperature was set to zero where supported (Gemini), Claude used its default thinking budget, and GPT-5.5 was run at the intermediate reasoning_effort setting (medium). The JSON-only output was natively enforced, where available (Gemini response_mime_type application/json, GPT-5.5 response_format json_object) and by prompt instruction for Claude. Each model received the same multipart input, a single text prompt followed by the 25 images in fixed order. The first call returned a structured report covering the histological type, Nottingham grade with component subscores, lymphovascular invasion (LVI), perineural invasion (PNI), ductal carcinoma in situ (DCIS) presence and pattern, extensive intraductal component (EIC), calcification, growth pattern, and stromal tumor-infiltrating lymphocytes. A second call submitted the same H&E tiles together with a text summary of the model’s own first-pass report, and the model predicted the molecular subtype (luminal, human epidermal growth factor receptor 2 [HER2]-positive, or triple-negative) based on the H&E morphology. Both prompts were identical across the three batches and were reproduced verbatim (Supplementary Material S1).

### Prompt engineering pre-investigation

Before the three-model comparison, we ran three pre-registered prompt-sensitivity pilots on an earlier Claude Sonnet 4.5 batch to determine whether targeted prompt redesign could move the model off its observed failure modes. Pilot version 2.1 (n=97; pre-registered 2026-04-29) tested active-search prompting for PNI against baseline absence-default in a 2×2 design of prompt language and tile budget (24 vs. 56 task tiles). Pilot version 2.2 (n=100; 2026-04-29) tested whether framing the invasive breast carcinoma of no special type (IBC-NST) as a diagnosis of exclusion and supplying per-special-type morphological criteria recovered special-type sensitivity. Pilot version 2.3 (n=100; 2026-05-13) tested whether replacing the directional grade language with hard quantitative thresholds, with or without additional tiles, shifted the observed central anchoring. The three pilots spanned 12 prompt variants and 4,056 API calls. Per-condition metrics, recognition versus classification decomposition, and test–retest reliability appear in the Supplement. Failure persisted across the tested range; therefore, a single fixed prompt was used in the main comparison.

### Reference standard

The reference was the clinical pathology report for the primary diagnostic specimen of each case signed by a board-certified breast pathologist (S-Y.J.) with >20 years of subspecialty experience. An automated rule-based parser extracted the histological type, with explicit handling of IBC-NST and the named special types, Nottingham score, and grade with the three component subscores, LVI, PNI, DCIS, EIC, calcification, growth pattern, and stromal tumor-infiltrating lymphocytes. Hormone receptor (HR) and HER2 status were obtained from institutional records, and surrogate molecular subtypes were assigned according to the St. Gallen 2013 criteria.^19^ For the analysis, the subtypes were collapsed into three treatment-relevant classes: luminal (HR-positive, HER2-negative), HER2-positive (any HER2-positive, regardless of HR), and triple-negative (HR-negative, HER2-negative).

### Statistical analyses

For each item, we compared the model output with the reference and reported the item-level concordance. For binary items, we reported raw concordance, sensitivity, specificity, and Cohen’s kappa together so that prevalence-driven inflation of raw agreement was visible. Histological type was summarized by exact concordance with class-wise sensitivity and Nottingham grade by linear- and quadratic-weighted kappa. The three models were compared for the same 427 paired cases using the McNemar test for binary items (exact binomial test when there were <25 discordant pairs, continuity-corrected chi-squared otherwise) and the Bowker symmetry test for the grade confusion matrix. P-values were two-sided and unadjusted, given the small number of prespecified comparisons. Ninety-five percent confidence intervals (CIs) for kappa were obtained from a nonparametric bootstrap with 1,000 resamples (seed 42). Subtype concordance was determined using broad kappa and per-class sensitivity in 414 evaluable cases. Two supplementary 28-case analyses compared GPT-5.5 at high versus medium reasoning_effort and Claude Opus 4.8 versus Sonnet 4.6 using the per-item McNemar test.

### Ethics

This study was approved by the Institutional Review Board (IRB) of the Catholic Medical Center (IRB no. OC26RISI0052), which waived the requirement for informed consent given the retrospective design and use of anonymized data. Identifiers were removed during slide selection.

## Results

### Cohort

Of the 427 analyzed cases, 391 had a reference Nottingham grade, 414 had a classifiable IHC subtype, and 427 had a reference histological type. The reference grade distribution was grade 1 in 53, grade 2 in 212 (54%), and grade 3 in 126 cases. The reference subtype distribution was luminal in 193, HER2-positive in 165, and triple-negative breast cancer (TNBC) in 56 cases.

### Histological type

By raw exact concordance among the 372 cases with an assignable reference histological type, Claude Sonnet 4.6 ranked highest at 87.4% (325 of 372), compared with 66.1% (246 of 372, Gemini 2.5 Pro) and 56.5% (210 of 372, GPT-5.5) (Table 1). This ranking did not survive the class-wise analysis. Of 23 invasive lobular carcinomas, Claude correctly classified 0 (sensitivity, 0.0%; 95% binomial CI, 0.0 to 14.3%), Gemini 9 (39.1%), and GPT-5.5 1 (4.3%); of the seven invasive micropapillary carcinomas, Claude classified 0 (0.0%), Gemini 1 (14.3%), and GPT-5.5 2 (28.6%) (Figure 2). Claude assigned the IBC-NST to every special-type case in the cohort, including all 23 lobular, all seven micropapillary, and the papillary, mucinous, and tubular cases. Therefore, the model with the highest raw concordance showed the lowest sensitivity for non-IBC-NST histology.

**Table 1.** Three-way item-level concordance against the pathologist reference. Exact concordance is shown for histologic type and Nottingham grade; sensitivity, specificity, and Cohen’s kappa (95% bootstrap CI) for the binary items; and quadratic-weighted kappa

| Item | Metric | Claude Sonnet 4.6 | Gemini 2.5 Pro | GPT-5.5 |
| --- | --- | --- | --- | --- |
| Histologic type | Exact concordance | 0.874 | 0.661 | 0.565 |
| Nottingham grade | Exact concordance | 0.552 | 0.528 | 0.340 |
| Nottingham grade | Quadratic-weighted kappa (95% CI) | 0.279 (0.203 to 0.354) | 0.411 (0.329 to 0.487) | 0.251 (0.194 to 0.310) |
| LVI | Sensitivity / specificity | 0.02 / 0.98 | 0.31 / 0.74 | 0.00 / 1.00 |
| LVI | Kappa (95% CI) | 0.005 (-0.057 to 0.108) | 0.050 (-0.052 to 0.153) | 0.000 |
| PNI | Sensitivity / specificity | 0.00 / 1.00 | 0.12 / 0.96 | 0.00 / 1.00 |
| PNI | Kappa | 0.000 | 0.084 | 0.000 |
| DCIS | Sensitivity / specificity | 1.00 / 0.00 | 0.72 / 0.47 | 0.48 / 0.80 |
| DCIS | Kappa (95% CI) | 0.000 | 0.125 (0.012 to 0.228) | 0.124 (0.050 to 0.198) |
| EIC | Kappa (95% CI) | 0.000 | 0.338 (0.207 to 0.466) | 0.104 (0.027 to 0.199) |
| Calcification | Kappa (95% CI) | -0.027 (-0.185 to 0.142) | 0.069 (-0.008 to 0.155) | 0.192 (0.093 to 0.288) |

LVI, lymphovascular invasion; PNI, perineural invasion; DCIS, ductal carcinoma in situ; EIC, extensive intraductal component; CI, confidence interval. Evaluable n differs by item and model (cases not assessable by the model or not reported in the reference are excluded): histologic type 372/372/372; Nottingham grade 391/390/376; LVI 182/399/399; PNI 404/404/404; DCIS 330/299/245; EIC 332/223/240; calcification 107/386/292 (Claude / Gemini / GPT-5.5). Kappa 95% CIs are from a 1,000-resample bootstrap.

**Figure 2.**
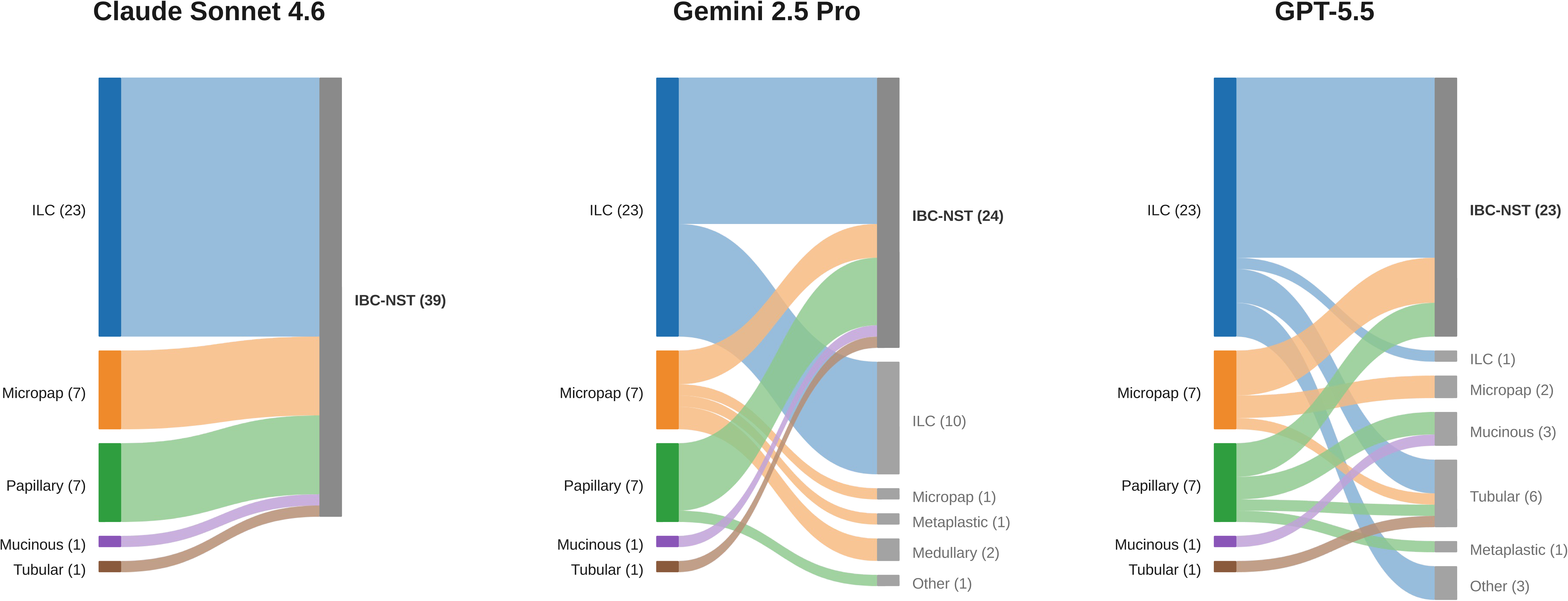
Fate of special-type carcinomas by model (n=39 special-type cases) Left, reference special-type histology (with n); right, model-assigned type.

### Nottingham grade

The exact grade concordance rates were 55.2% (Claude), 52.8% (Gemini), and 34.0% (GPT-5.5). Against a grade 2 majority reference, each model concentrated its output on a different grade. Among cases with an assignable grade, Claude assigned grade 2 in 292 of 391 (74.7%), Gemini assigned grade 3 in 215 of 390 (55.1%), and GPT-5.5 assigned grade 1 in 209 of 376 (55.6%) cases (Figure 3). The quadratic-weighted kappa values were 0.279 (Claude, 95% CI 0.203 to 0.354), 0.411 (Gemini, 0.329 to 0.487), and 0.251 (GPT-5.5, 0.194 to 0.310); none of the models reached the threshold for substantial agreement (kappa, at least 0.61).

**Figure 3.**
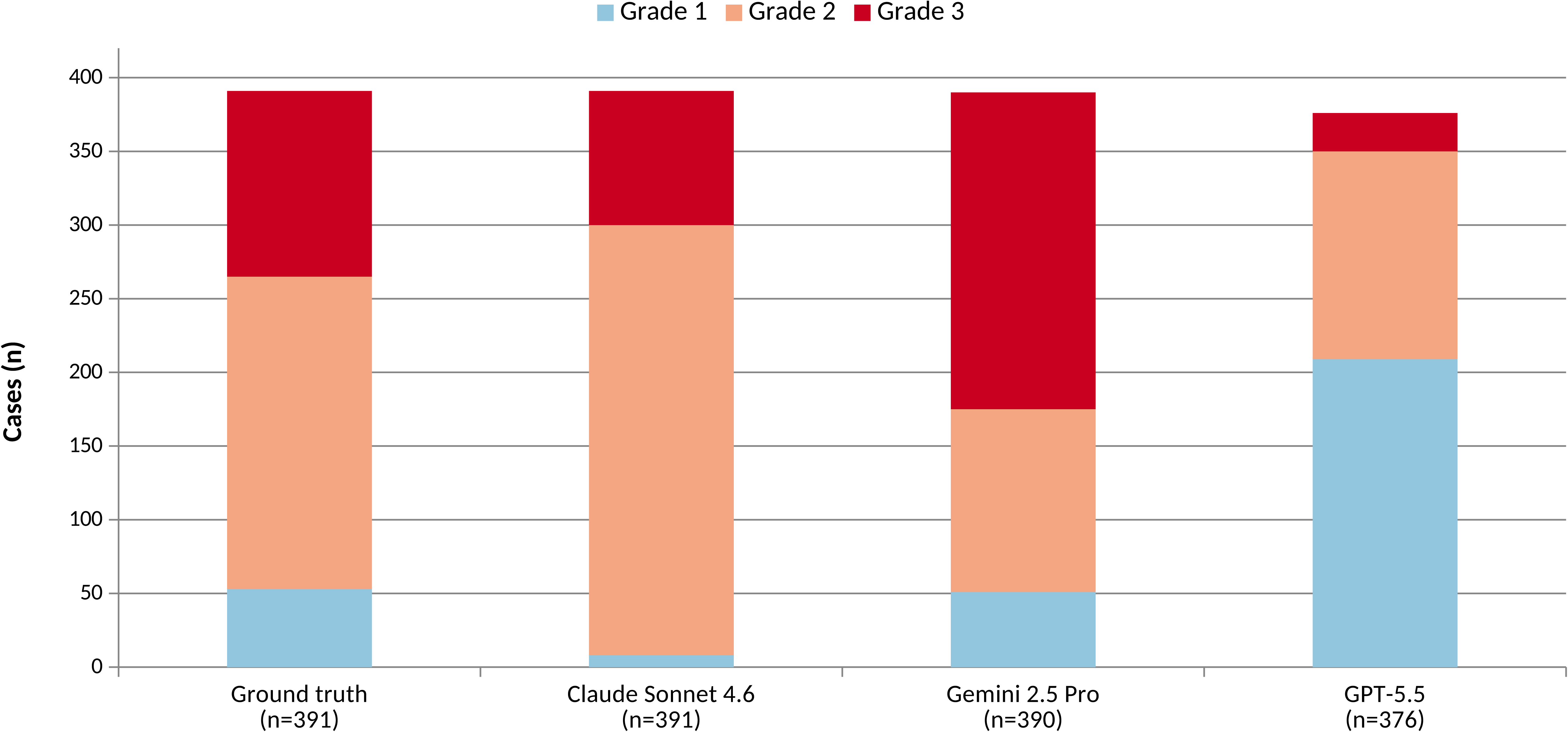
Nottingham grade distribution – three different anchoring modes. Reference modal grade is grade 2; Claude anchors to grade 2, Gemini to grade 3, and GPT-5.5 to grade 1. Counts on grade-evaluable cases.

### Lymphovascular and perineural invasion

LVI sensitivities were 0.02 (Claude), 0.31 (Gemini), and 0.00 (GPT-5.5), with specificities of 0.98, 0.74, and 1.00, respectively; the LVI kappa values were 0.005 (95% CI, –0.057 to 0.108), 0.050, and 0.000, respectively (Table 1). PNI sensitivities were 0.00 (Claude), 0.12 (Gemini), and 0.00 (GPT-5.5). Claude and GPT-5.5 reported these findings as absent in nearly all cases, whereas Gemini reported them as present more often, at the cost of specificity.

### In situ disease and other binary items

The DCIS-presence sensitivity and specificity were 1.00 and 0.00 (Claude), 0.72 and 0.47 (Gemini), and 0.48 and 0.80 (GPT-5.5), respectively; the DCIS kappa values were 0.000, 0.125, and 0.124, respectively. The EIC kappa values were 0.000 (Claude), 0.338 (Gemini, 95% CI 0.207 to 0.466), and 0.104 (GPT-5.5). The calcification kappa values were –0.027 (Claude), 0.069 (Gemini), and 0.192 (GPT-5.5) (Table 1). Across the binary item set, the only chance-corrected agreement with a bootstrap CI bounded away from zero by a margin consistent with fair agreement was the Gemini EIC (Figure 4). For calcification, among the cases in which a model was committed to a binary call, the conditional rates of a present call were 82% for Claude, 26% for Gemini, and 28% for GPT-5.5 (Supplementary Figure).

**Figure 4.**
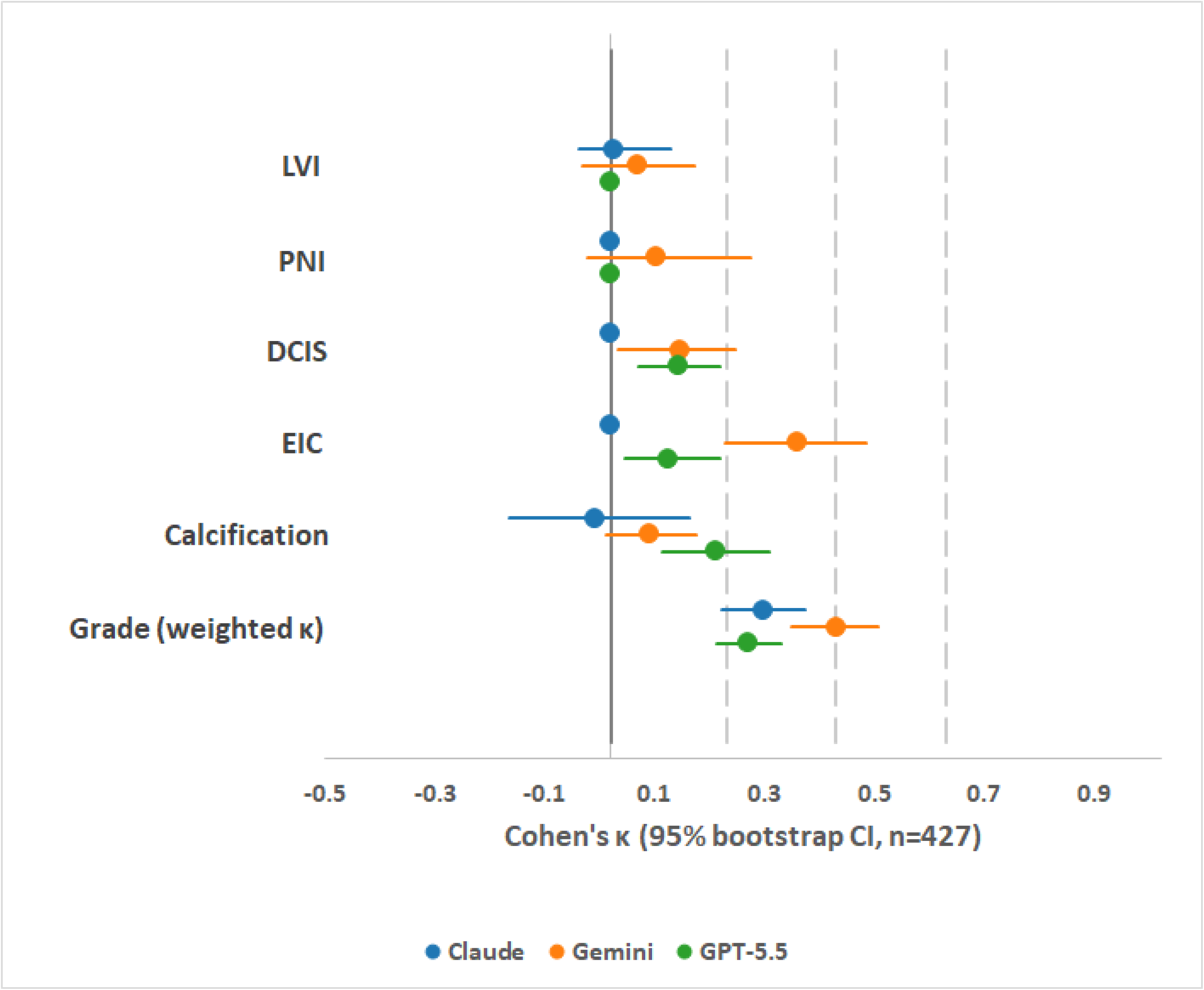
Cohen’s kappa for binary items and Nottingham grade, by model. Points are item-level Cohen’s kappa for the binary findings (LVI, PNI, DCIS, EIC, calcification) and quadratic-weighted kappa for Nottingham grade, each with 95% bootstrap confidence intervals (n=427); the vertical line marks kappa = 0. Evaluable n varies by item and model (Table 1).

### Molecular subtype

Subtypes were assessed in 414 cases using a three-class scheme. The luminal sensitivity was high for all three models (97.4% Claude, 83.9% Gemini, 90.7% GPT-5.5). The HER2-positive sensitivity was low for all three subtypes (n=165): 1.8% (Claude, 3 of 165), 3.6% (Gemini, 6 of 165), and 1.2% (GPT-5.5, 2 of 165). TNBC sensitivity diverged (n=56): 0% (Claude, 0 of 56), 48.2% (Gemini, 27 of 56), and 17.9% (GPT-5.5, 10 of 56). Gemini returned 97 TNBC calls against a TNBC prevalence of 56 (positive predictive value, 27.8%), whereas Claude assigned all TNBC cases to luminal categories. The broad three-class kappa values were 0.012 (Claude), 0.156 (Gemini), and 0.072 (GPT-5.5).

### Pairwise model differences

Pairwise McNemar tests showed significant differences for LVI across all three vendor pairs (all p<0.001): for PNI (Claude vs. Gemini and Gemini vs. GPT-5.5, both p=0.004; Claude vs. GPT-5.5 p=1.0, reflecting identical absence-default outputs); for DCIS (all three pairs p<0.001); for EIC (Claude vs. Gemini and Claude vs. GPT-5.5, both p<0.001; Gemini vs. GPT-5.5, p=0.57); and for calcification (Claude vs. Gemini and Claude vs. GPT-5.5, both p<0.001; Gemini vs. GPT-5.5, p=0.012) (Table 2). Bowker symmetry tests for the Nottingham grade were significant for all three pairs (all p<0.0001).

**Table 2.** Pairwise model comparisons. McNemar two-sided p-values for each binary item across the three vendor pairs; Bowker symmetry test (chi-square, df=3) for the Nottingham grade confusion matrix.

| Item | Claude vs Gemini | Claude vs GPT-5.5 | Gemini vs GPT-5.5 |
| --- | --- | --- | --- |
| LVI (McNemar) | <0.001 | <0.001 | <0.001 |
| PNI (McNemar) | 0.004 | 1.0 | 0.004 |
| DCIS (McNemar) | <0.001 | <0.001 | <0.001 |
| EIC (McNemar) | <0.001 | <0.001 | 0.57 |
| Calcification (McNemar) | <0.001 | <0.001 | 0.012 |
| Nottingham grade (Bowker chi-square) | 148.9 (p<0.0001) | 241.2 (p<0.0001) | 268.2 (p<0.0001) |

LVI, lymphovascular invasion; PNI, perineural invasion; DCIS, ductal carcinoma in situ; EIC, extensive intraductal component.

### Prompt-sensitivity pilots and supplementary analyses

Across the 12 prompt variants and 4,056 API calls in the three pre-registered pilots, none of the variants achieved nonzero sensitivity for PNI (0 of 17 positive cases under all v2.1 conditions) or special-type recognition (0 of 40 under all v2.2 conditions). In all 40 special-type cases under the most directed v2.2 prompt, the model named the diagnostic morphological features in its free-text description while still outputting the IBC-NST as the structured classification. In v2.3, a hard-threshold grade prompt raised the grade 3 sensitivity from 0.500 to 0.950 but lowered the quadratic-weighted kappa from 0.336 to 0.273. In the supplementary 28-case analyses, raising GPT-5.5 reasoning_effort from medium to high produced no significant change in any binary item (all McNemar p=1.0), with an LVI sensitivity 0 of 28 retained. Replacing Claude Sonnet 4.6 with Claude Opus 4.8 shifted the DCIS and LVI call distributions (McNemar p<0.001 and p=0.02), but LVI sensitivity remained 0 of 28 and special-type recognition 1 of 28.

## Discussion

In this study of three flagship commercial MLLMs on identical H&E images and identical prompts in 427 invasive breast cancers, the models produced systematically different diagnoses across all treatment-determining features examined. These three findings define the results. First, the raw concordance and chance-corrected agreements ranked the models in the opposite order. Second, each vendor failed in a fixed and clinically distinct manner. Third, no model reached a chance-corrected agreement approaching the published inter-pathologist levels for any item.

Because the images, tiles, and prompt were held constant, the model vendor was the principal variable under experimental control. However, the rank order of the models reversed depending on the metric. Claude’s 87.4% histologic-type concordance coincided with zero sensitivity for every special type, because the model assigned IBC-NST to all three, and the cohort was dominated by IBC-NST. Raw concordance rewarded majority-class collapse and penalized the models that attempted, imperfectly, to recover special types; chance correction reversed the ordering. This is a known property of agreement statistics with skewed prevalence, in which a high observed agreement coexists with a near-zero kappa.^20,21^ A deployment decision based on raw concordance alone would have identified the least discriminating model as the best. Because most breast pathology items have a skewed prevalence, chance-corrected agreement should be reported alongside raw concordance.

The direction of error differed by vendor and corresponded to opposite clinical consequences. For LVI, Claude and GPT-5.5 returned a negative call in nearly all positive cases (false negatives), tending toward understaging and omission of adjuvant therapy, whereas Gemini returned more positive calls at low specificity (false positives), tending toward overtreatment. The same contrast appeared for TNBC: Claude assigned every case to a non-TNBC class, whereas Gemini overcalled TNBC, with fewer than one in three of its TNBC calls correct. Overall, no vendor was free of directional misclassification.

These patterns are more consistent with current commercial MLLMs that behave as fixed-bias decision systems rather than as additional readers whose errors are random around a shared truth. Pre-investigation pilot studies supported this interpretation. Twelve prompt variants across the three failure modes did not correct the failure in the targeted domain. In pilot v2.2, in all 40 special-type cases under the most directed prompt, the model named the diagnostic morphological features in free text, but still output IBC-NST as the classification. Therefore, the failure lies in the model’s prior classification prior, not in visual perception. Hard-threshold prompting in v2.3 redirected the grade anchoring without improving the quadratic-weighted kappa, and neither reasoning-effort escalation nor an intra-vendor version upgrade altered the failure direction. Within the range tested, the output reflects a vendor-specific prior that prompt engineering can redirect, but not eliminate.

Published inter-pathologist agreement provides a reference for how far these results sit below the human reading they would replace. The interobserver weighted kappa for the Nottingham grade ranges from 0.5 to 0.7.^22^ The model’s quadratic-weighted kappa for the grade fell below this range for all three vendors; the best single value, the Gemini grade (0.411), remained below the lower bound. For binary items, the model kappa values were clustered near zero, indicating a near-chance agreement. Therefore, the conclusion that no model reaches inter-pathologist agreement holds, even when the single-rater reference is judged against the published multi-rater agreement.

The failure pattern is consistent with the gap between general-purpose multimodal models and domain-trained computational pathology. Task-specific deep learning pipelines^12,23–26^ and pathology foundation models pretrained on at least 1 million slides^13–15^ report benchmark-level performance for breast Nottingham grade and receptor status from H&E, and vision-language assistants built on pathology data outperform general-purpose multimodal models on pathology benchmarks.^16^ The present results quantify the gap for treatment-determining breast pathology items and locate them in the models’ classification priors rather than in image access. Under the most directed special-type prompt, the models described the correct morphology but still returned the IBC-NST. Invasive carcinoma of no special type is defined by the WHO classification as a diagnosis of exclusion.^4,27^ Thus, special-type recognition requires a learned prior that is sufficiently strong to override the dominant class. The single advantage of the API-only configuration is access, because it requires no local GPU, model hosting, or domain fine-tuning; the cost of that access is the performance ceiling documented here. Because the models perceived the morphology but defaulted to a common class, future use will depend on giving them domain-specific reference information rather than rewording the prompt: labeled example images in the prompt, automatic retrieval of similar reference cases, or handoff to a pathology-specific model. Each depends on balanced, IHC-confirmed H&E datasets for the special types and receptor subtypes that failed here, and each sacrifices part of the low-barrier simplicity of API-only access, a tradeoff that should be tested prospectively.

The clinical role supported by these data is assistive rather than autonomous. At an input- and output-token cost on the order of USD 0.20–0.50 per case, a commercial MLLM could lower the barrier to a structured first-pass report in settings where pathologist throughput, rather than the absence of any reading, is the binding constraint, including the low- and middle-income contexts that motivated this study. This role presupposes whole-slide digitization, as the model reads scanned images, and, for receptor subtype, IHC, which the model cannot recover from H&E. Both are uneven in such settings, and a survey of population-based pathology centers in sub-Saharan Africa reported in-house IHC in approximately half of those surveyed.^28^ Where these foundations are in place but specialist time is the limiting factor, an MLLM-generated first-pass report could help extend the reach of scarce pathologists. The present results bound that role: An output that underdetects or overcalls in a fixed direction cannot stand as a primary read, and any use would require a pathologist to retain the diagnosis and apply vendor-aware caveats in the direction of error. A constraint specific to this route is that patient images are transmitted to a commercial, often cross-border, server subject to data protection rules.

This study has some limitations. The cohort is from a single university-affiliated tertiary hospital in the Republic of Korea, not from a low- or middle-income setting; thus, generalizability to the populations concerned with the access argument is untested and requires prospective evaluation. The reference standard was the sign-out diagnosis of a single subspecialty breast pathologist. Within-study interobserver agreement could not be calculated, and the kappa values reported here were not directly comparable with the multi-rater benchmarks. Because molecular subtypes are defined by IHC, H&E-only input sets a ceiling for subtype inference. Each model was evaluated at a single dated version; model versioning is rapid, and although the supplementary analyses of GPT-5.5 reasoning effort and Claude Opus 4.8 shifted call distributions without altering the failure direction, transfer to other releases is not guaranteed. The tile-sampling protocol (one thumbnail and 24 tiles across three magnifications) does not exhaust slide content and may miss focal findings, such as mitotic hot spots and small lymphovascular foci. The model also received tiles from a single representative slide, whereas the reference diagnosis drew on all slides, IHC, and clinical contexts, reflecting the asymmetry of the available information. This study did not include a formal cost-effectiveness analysis or prospective deployment evaluation.

No current flagship commercial MLLM reached deployment-ready accuracy for any treatment-determining feature of the breast pathology examined in this study. Performance ranked differently under raw and chance-corrected agreements, and each vendor failed in its own fixed direction. Therefore, changing vendors altered the type of diagnostic error rather than removing it. These patterns persisted across 12 prompt variants, a reasoning effort escalation, and an intravendor version upgrade, indicating a vendor-specific prior rather than a prompt engineering artifact. The role supported by these data is assistive, as a supervised generator of draft structured reports that leave the diagnosis to the pathologist, not an autonomous reading. Whether supplying a missing domain prior can increase the performance ceiling without losing the low-barrier access that defines these tools is a question for prospective studies.

## Declarations

### Ethics approval

The study was approved by the Institutional Review Board of the Catholic Medical Center (IRB no. OC26RISI0052), with a waiver for informed consent.

### Data availability

The WSIs and linked clinical pathology data contain potentially identifiable patient information and cannot be made publicly available. De-identified data underlying the analyses are available from the corresponding author upon reasonable request, subject to approval by the Institutional Review Board of the Catholic Medical Center and a data-use agreement.

### Code availability

The analysis code, including WSI tiling, item-level concordance analyses, agreement statistics, and paired McNemar and Bowker tests implemented in Python, will be made available upon publication at https://github.com// and archived at Zenodo (https://doi.org/10.5281/zenodo.). The repository does not contain patient data.

### Prompt availability

The prompts used in this study are provided in the Supplementary Material S1 (condensed version). Full original prompts are available from the corresponding author upon request.

### Author contributions

Y-J.K. conceived and designed the study, built the image-processing and analysis pipeline, performed the statistical analysis, generated the figures and tables, and wrote the manuscript. S-Y.J., a board-certified breast pathologist, provided the pathology reference standard, collected and verified the case data, advised on pathology content, and drafted the pathology components of the manuscript. S.K. assisted with data collection and study administration. All authors had access to the study data, contributed to interpretation, critically revised the manuscript, and approved the final version. Y-J.K. is the guarantor and had final responsibility for the decision to submit.

---

**Supplementary Figure.**
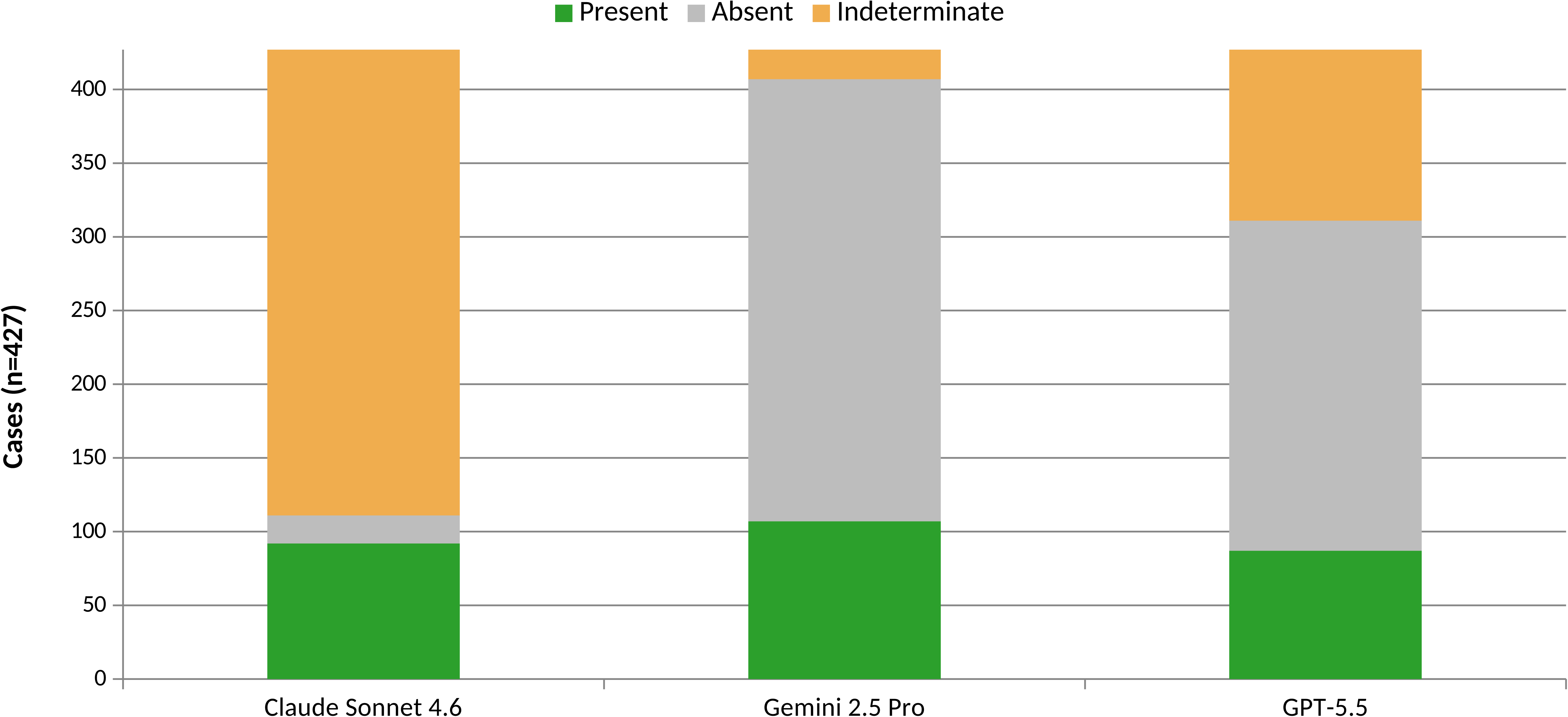
Calcification call distribution by model

